# Comparisons of healthcare resource utilisation and costs between Brugada syndrome and congenital long QT syndrome: a territory-wide study

**DOI:** 10.1101/2022.11.12.22282257

**Authors:** Sharen Lee, Cheuk To Chung, Danny Radford, Oscar Hou In Chou, Teddy Tai Loy Lee, Keith Sai Kit Leung, Leonardo Roever, Rajesh Rajan, George Bazoukis, Konstantinos P Letsas, Shaoying Zeng, Fang Zhou Liu, Wing Tak Wong, Tong Liu, Gary Tse

## Abstract

**Introduction:** Healthcare resource utilisation and costs are important metrics of healthcare burden, but they have rarely been explored in the setting of cardiac ion channelopathies.

**Purpose:** The aim of this study is to compare HCRUs and costs between patients with Brugada syndrome (BrS) and congenital long QT syndrome (LQTS) in a single city of China.

**Methods:** This was a territory-wide retrospective cohort study of consecutive BrS and LQTS patients at public hospitals or clinics in Hong Kong, China. HCRUs and costs (in USD) for accident and emergency (A&E), inpatient, general outpatient and specialist outpatient attendances were analysed over a 19-year period (2001-2019) at the cohort level. Comparisons were made between BrS and LQTS cohorts using incidence rate ratios (IRRs [95% confidence intervals]).

**Results:** Over the 19-year study period, 516 BrS (median age of initial presentation: 51 [interquartile range: 38-61] years, 92% male) and 134 LQTS (median age of initial presentation: 21 [9-44] years, 32% male) patients were included. BrS patients had lower total costs compared to LQTS patients (2,008,126 [2,007,622-2,008,629] vs. 2,343,864 [2,342,828-2,344,900]; IRR: 0.857 [0.855-0.858]). For specific attendance types, BrS patients had higher costs for A&E attendances (83,113 [83,048-83,177] vs. 70,604 [70,487-70,721]; IRR: 1.177 [1.165-1.189]) and general outpatient services (2,176 [2,166-2,187] vs. 921 [908-935]; IRR: 2.363 [2.187-2.552]). However, they had lower costs for inpatient stay (1,391,624 [1,391,359-1,391,889] vs. 1,713,742 [1,713,166-1,714,319]; IRR: 0.812 [0.810-0.814]) and to a smaller extent, lower costs for specialist outpatient services (531213 [531049-531376] vs. 558597 [558268-558926]; IRR: 0.951 [0.947-0.9550]) compared to LQTS patients.

**Conclusion:** Overall, BrS patients consume 14% less healthcare resources compared to LQTS patients in terms of attendance costs. BrS patients require more A&E and general outpatient services, but less inpatient and specialist outpatient services than LQTS patients. Further studies are needed to examine patient-based attendances and costs to identify subgroups of high HCRU users for both cohorts.

## Introduction

Cardiac channelopathies can be categorized by the development of arrhythmias due to abnormalities in the function and/or structure of ion channels, resulting in syncope and sudden cardiac death (SCD) (1). In recent years, there has been rising interest regarding the management of Long QT Syndrome (LQTS) and Brugada Syndrome (BrS). Both conditions involve mutations in the *SCN5A* gene which encodes for the pore-forming subunit of the cardiac sodium ion channel (2). LQTS is a relatively well-documented cardiac condition, with more than 15 disease-causing genes identified and may be seen in approximately 0.1% of the general population (3). In contrast, the interpretation of genetic variants in BrS is difficult, with approximately 25% of cases attributed to SCN5A mutations (4). Therefore, this presents a greater challenge in risk stratification and clinical management of BrS (5-7).

However, there is little understanding of the healthcare burden of LQTS and BrS patients. The provision of genetic testing options, implantable cardioverter-defibrillators (ICD), hospital admissions from arrhythmia-related symptoms, and the need for specialist outpatient services follow-up for device and arrhythmia management and monitoring are major drivers for healthcare expenditure (8, 9). As of now, little research has been dedicated to investigating the healthcare resource utilization (HCRU) and related costs in the setting of cardiac ion channelopathies. With increasing awareness and diagnosis of both conditions, there may be a subsequent increase in service demand, thus raising the concern for prioritization in healthcare interventions and specific cost-effectiveness estimations. Without a comprehensive analysis of the cost-effectiveness of medical technologies, this may undermine the benefit of healthcare policies. Hence, the aim of this study is to compare the healthcare resource utilization (HCRU) and related costs between BrS and LQTS patients in Hong Kong, China.

## Methods

### Study population

The study was part of a wider study on cardiac arrhythmias approved by The Joint Chinese University of Hong Kong-New Territories East Cluster Clinical Research Ethics Committee.This territory-wide retrospective cohort study includes patients diagnosed with BrS or LQTS between the 1^st^ of January, 1997 to the 31^st^ of December, 2020 in public hospitals or clinics in Hong Kong. Centralised electronic health records from the Clinical Data Analysis and Reporting System (CDARS) were evaluated for patient identification and data extraction. This system has been used previously by our team and other teams for healthcare resource utilisation and cost analysis for catecholaminergic polymorphic ventricular tachycardia (CPVT) (10), cancer patients receiving immunotherapy (11), and COVID-19 (12, 13). The diagnosis of LQTS and BrS was made initially by case physicians and was further verified by G.T. through documented ECGs, case notes, genetic reports and diagnostic test results in accordance with the 2017 Expert Consensus Statement for BrS (14).

### Clinical and Electrocardiographic Data Collection

Our team has published previously using these LQTS and BrS cohorts for risk prediction (15). Baseline clinical data was extracted from the electronic health records. This included: (1) sex; (2) age of first characteristic ECG presentation and last follow-up; (3) follow-up duration; (4) syncope manifestation and its frequency; (5) family history of SCD and the specific ion channelopathy; (6) performance of electrophysiological study (EPS), 24-hours Holter study, ion channelopathy-specific genetic testing of the RYR2 gene, and the respective results; (7) presentation of sustained VT/VF and its frequency; (8) presence of other arrhythmias; (9) implantation of ICD; (10) ECG performance; (11) period between the initial presentation of characteristic ECG and the first post-diagnosis VT/VF episode; (12) initial disease manifestation (asymptomatic, syncope, VT/VF); (13) occurrence, cause and age of death. The baseline ECG was extracted at the earliest time possible after the presentation of an initial characteristic ECG pattern.

### Statistical, Healthcare Resource Utilisation and Cost Analyses

Categorical variables were represented as a total sum and percentage. Continuous and discrete variables were expressed as a mean and standard deviation (SD) value. The HCRU and costs for accident and emergency (A&E), inpatient, general outpatient and specialist outpatient attendances were analysed over a 19-year period (2001-2019). Incidence rate ratios (IRRs [95% confidence intervals]) were used to conduct comparisons between the BrS and LQTS cohort. The attendance costs were calculated using unit costs in reference to the standard of the local government. Final cost values were presented in USD. Statistical significance was defined as p<0.05. All statistical analysis was performed using R Studio (Version: 1.3.1073).

## Results

### Baseline Characteristics

In this study, 516 BrS patients and 134 LQTS patients were included. The average age at first presentation was much younger for the LQTS cohort compared to the BrS cohort (27.6 ± 23.8 vs. 49.9 ± 16.2). In addition, the LQTS cohort had a greater percentage of females (67.9% vs. 7.6%), as well as more patients with a family history of the disease (43.3% vs. 3.1%) and VF/SCD (14.9% vs. 7.9%) compared to the BrS cohort. The number of genetic tests performed was also higher in the LQTS cohort compared to the BrS cohort (84 vs. 51). Interestingly, the BrS cohort performed significantly more EPS (112 vs. 6) and had a greater proportion of induced VT/VF (14.7% vs. 3.0%). In regards to the baseline ECG characteristics, BrS patients had overall longer PR interval (169.5 ± 29.0 vs. 161.8 ± 29.8) and P-wave duration (114.7 ± 18.1 vs. 105.1 ± 17.5) but shorter QTc interval (368.9 ± 42.4 vs. 488.5 ± 44.4) compared to the LQTS cohort. The baseline characteristics comparing the LQTS and BrS cohort are summarised in **Table 1**.

**Table 1.** Baseline characteristics of the study cohort. Categorical and continuous variables were compared between LQTS and BrS patients.

| Variable | LQTS (n=134) | BrS (n=516) |
| --- | --- | --- |
| **Clinical characteristics** |  |  |
| Female | 91 (67.9) | 39 (7.6) |
| Age at first presentation | 27.6 ± 23.8 | 49.9 ± 16.2 |
| Family history of LQTS/BrS | 58 (43.3) | 16 (3.1) |
| Family history of VF/SCD | 20 (14.9) | 41 (7.9) |
| Syncope | 68 (54.4) | 222 (43.0) |
| Spontaneous VT/VF in follow-up | 51 (38.1) | 80 (15.5) |
| Initial VT/VF | 34 (25.4) | 42 (8.1) |
| Treadmill performed | 50 (37.3) | 63(12.2) |
| EPS | 6 (4.5) | 112(21.7) |
| *Induced VT/VF under EPS* | 4 (3.0) | 76 (14.7) |
| ICD | 51 (38.1) | 136 (26.4) |
| Genetic test | 84 (62.7) | 51 (9.9) |
| **Baseline ECG characteristics** |  |  |
| Heart rate (bpm) | 76.7 ± 23.4 | 80.9 ± 20.0 |
| P-wave duration (ms) | 105.1 ± 17.5 | 114.7 ± 18.1 |
| PR interval (ms) | 161.8 ± 29.8 | 169.5 ± 29.0 |
| QRS interval (ms) | 95.4 ± 21.9 | 106.4 ± 22.7 |
| QT interval (ms) | 444.2 ± 71.8 | 416.5 ± 33.2 |
| QTc Interval (ms) | 488.5 ± 44.4 | 368.9 ± 42.4 |
| P axis | 54.9 ± 40.4 | 61.3 ± 22.3 |
| QRS axis | 55.1 ± 56.9 | 58.7 ± 39.6 |
| T axis | 52.9 ± 54.9 | 54.3 ± 26.0 |
| R wave in lead V5 | 1.2 ± 0.7 | 1.5 ± 0.6 |
| S wave in lead V1 | 0.7 ± 0.4 | 0.6 ± 0.3 |

### Healthcare Resource Utilisation and Cost Analysis

The total number of attendances for A&E, inpatient, general and specialist outpatient setting in the cohort is as follows: 5154, 4140, 373 and 34049 for the BrS cohort and 1137, 1285, 41, 9298 for the LQTS cohort. Both cohorts demonstrated the highest number of attendance in the specialist outpatient settings, however the BrS cohort had a greater overall number of attendance compared to the LQTS cohort (43716 vs. 11761). In addition, the attendance number of inpatient length of stays of the BrS cohort were also significantly higher than the LQTS cohort (20813 vs. 6656). The attendance and costs of the BrS and LQTS cohort are shown in **Table 2**.

**Table 2.**
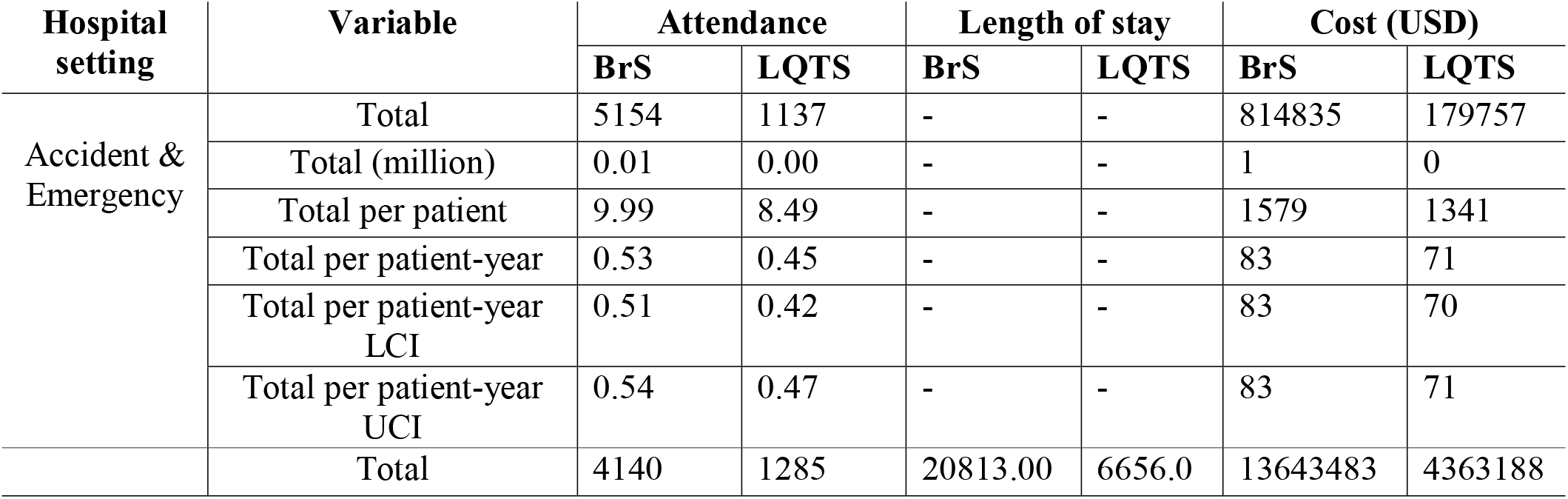
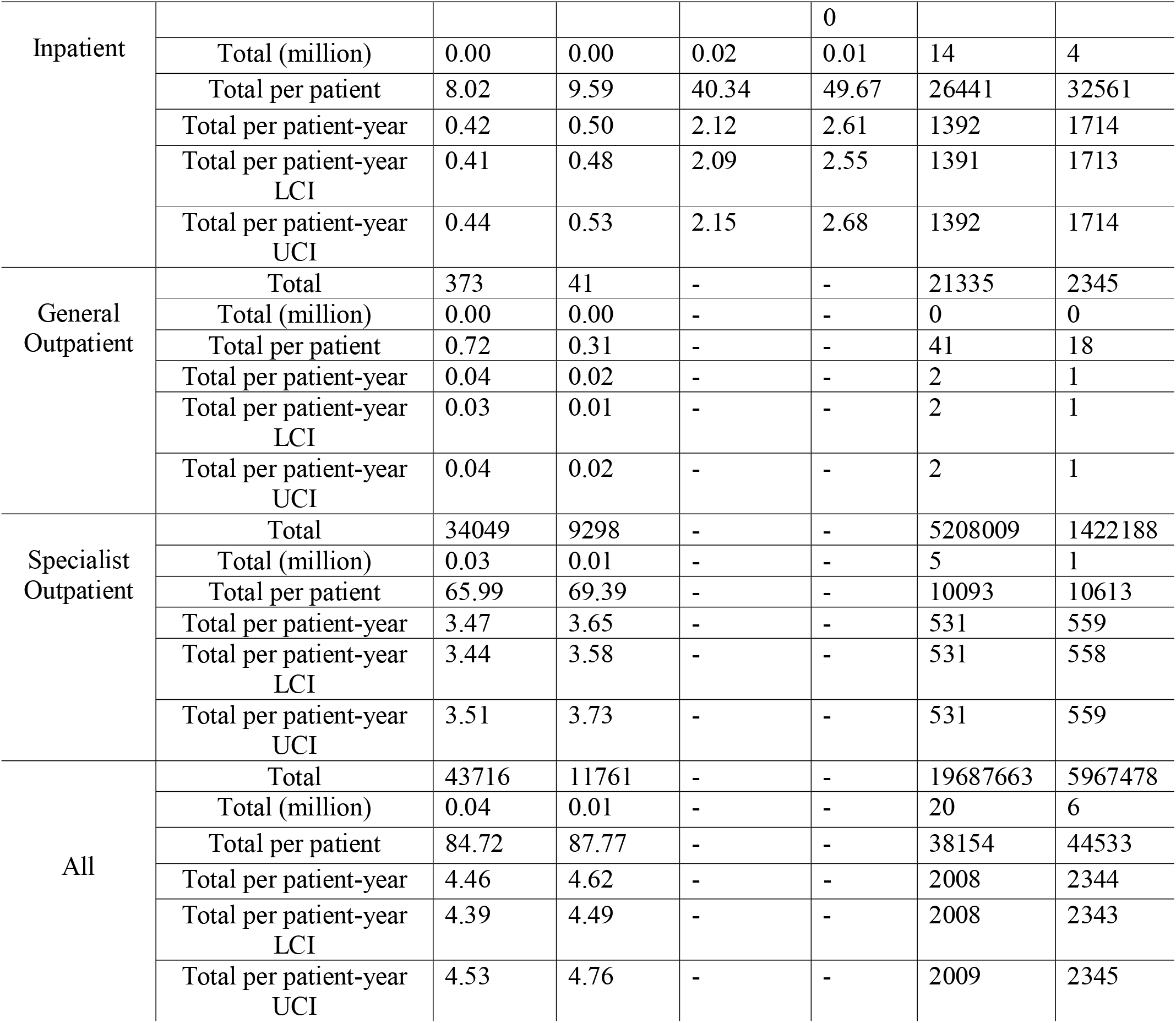
Cohort-level attendance, length of stay and costs for BrS and LQTS patients. Median (lower and upper 95% confidence intervals) values are presented. Costs shown are in US dollars.

In comparison to LQTS patients, BrS patients had lower overall costs (2,008,126 [2,007,622-2,008,629] vs. 2,343,864 [2,342,828-2,344,900]; IRR: 0.857 [0.855-0.858]) (Table 3). To corroborate BrS patients had higher costs for A&E attendances (83,113 [83,048-83,177] vs. 70,604 [70,487-70,721]; IRR: 1.177 [1.165-1.189]) and general outpatient services (2,176 [2,166-2,187] vs. 921 [908-935]; IRR: 2.363 [2.187-2.552]) relative to LQTS patients. In contrast, LQTS patients had higher costs for inpatient stay (1,713,742 [1,713,166-1,714,319] vs. 1,391,624 [1,391,359-1,391,889]; IRR: 0.812 [0.810-0.814]) and slightly higher costs for specialist outpatient services (558597 [558268-558926]; vs. 531213 [531049-531376]; IRR: 0.951 [0.947-0.9550]) compared to BrS patients.

**Table 3.**
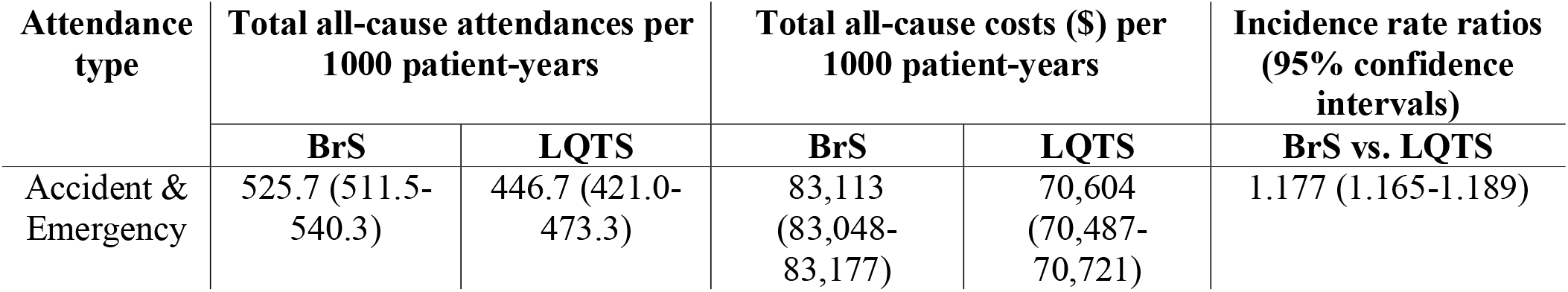
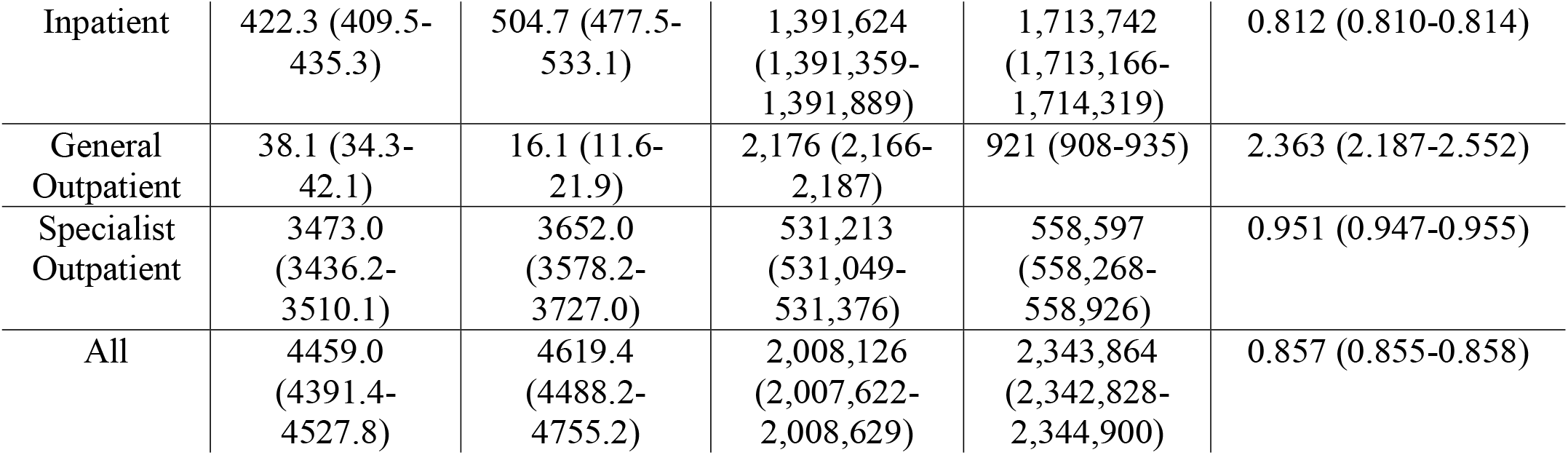
Cohort-level healthcare utilisation and costs for BrS and LQTS patients. Median (lower and upper 95% confidence intervals) values are presented. Costs shown are in US dollars.

| Attendance type | Total all-cause attendances per 1000 patient-years | | Total all-cause costs (\$) per 1000 patient-years | | Incidence rate ratios (95% confidence intervals) |
| --- | --- | --- | --- | --- | --- |
|  | BrS | LQTS | BrS | LQTS | BrS vs. LQTS |
| Accident & Emergency | 525.7 (511.5-540.3) | 446.7 (421.0-473.3) | 83,113 (83,048-83,177) | 70,604 (70,487-70,721) | 1.177 (1.165-1.189) |
| Inpatient | 422.3 (409.5-435.3) | 504.7 (477.5-533.1) | 1,391,624<br>(1,391,359-1,391,889) | 1,713,742<br>(1,713,166-1,714,319) | 0.812 (0.810-0.814) |
| General<br>Outpatient | 38.1 (34.3-42.1) | 16.1 (11.6-21.9) | 2,176 (2,166-2,187) | 921 (908-935) | 2.363 (2.187-2.552) |
| Specialist<br>Outpatient | 3473.0<br>(3436.2-3510.1) | 3652.0<br>(3578.2-3727.0) | 531,213<br>(531,049-531,376) | 558,597<br>(558,268-558,926) | 0.951 (0.947-0.955) |
| All | 4459.0<br>(4391.4-4527.8) | 4619.4<br>(4488.2-4755.2) | 2,008,126<br>(2,007,622-2,008,629) | 2,343,864<br>(2,342,828-2,344,900) | 0.857 (0.855-0.858) |

*Temporal trends of healthcare resource utilisation and costs from 2001 to 2019*

## Discussion

This is the first territory-wide cohort study in Hong Kong to compare the healthcare costs of LQTS and BrS. The major findings of this study is as follows: (1) BrS patients consume 14% less healthcare resources compared to LQTS patients; (2) BrS patients require more services from A&E and general outpatient setting; (3) LQTS patients require more services from inpatient and special outpatient setting.

The present study suggests that there are drastic differences in the healthcare burden of the two cardiac channelopathies. Due to the lower caseload of LQTS compared to BrS in Hong Kong (16-18), a greater percentage of patients had to undergo more cardiological examinations and consultations at a clinical genetics department. It may be argued that genetic testing plays a greater role in the diagnosis of LQTS relative to BrS because current genetic knowledge of LQTS is more advanced and the disease is more likely to have a genetic origin (19). Even for patients with inconclusive clinical scores, the current referral practice for LQTS often entails for aggressive treatment including rigorous restrictions on the patient’s lifestyle and primary ICD implantation (20). Resultantly, this may warrant unnecessary expenditure on patients who are at low-risk or have no risk of LQTS.

This notwithstanding, the family members of the LQTS patient are often included in the confirmatory testing process to screen for concealed or pre-clinical LQTS (21). Through early identification of familial LQTS, this will allow patients to receive timely secondary and tertiary prevention. Subsequently, this may lead to the increase in financial costs and anxiety for the patient. The 5-gene version of the FAMILION LQTS test costs approximately $5400 per index case and $900 per family member as a confirmatory test (22). Although newer technologies demonstrate great potential in reducing costs of intervention and detection of new mutations (23), the lack of competition minimizes the commercial incentive in finding new alternatives. However, current prices for diagnostic assessments were still significantly less expensive compared to previous genetic tests without genetic testing strategies (24). Despite advancements made in the understanding of LQTS genetics, the distinction between pathogenic and benign variants in LQTS-susceptibility genes remains challenging for physicians (25). Hence, this also warrants the need for further refinement of the clinical interpretation of LQTS to reduce the number of false positive and familial LQTS patients, and ultimately healthcare costs (26). Furthermore, it is also crucial to consider the relevant healthcare policies and insurance regulations of individual hospitals. Therefore, this may explain the healthcare cost discrepancies between the LQTS and BrS cohort.

### Strengths and limitations

Several major strengths were demonstrated in this study: (1) costs were estimated using standardized unit costs across extended follow-up periods; (2) the sampling of one of the largest cardiac channelopathies cohorts available enhances the reliability of study findings; (3) the use of a public, comprehensive electronic health record system from the city, incorporating attendances from 43 hospitals and their associated outpatient and ambulatory care facilities.

Several limitations should also be noted. The retrospective observational nature of this study suggests that results may be prone to coding errors, under-coding or missing data, resulting in information and selection bias. However, as the majority of patients were closely followed-up through annual consultations, the bias was amended with detailed follow-up and patient documentation. Although the database used already documents one of the largest cohorts of cardiac channelopathies in Asia, the sample size is still small compared to other cardiac diseases, especially the LQTS cohort. Consequently, this limits the validity of study findings. This is due to the fact that the prevalence of BrS and LQTS is low relative to other cardiac diseases in Hong Kong. It is prudent to recognize that our cost analyses require additional external validation in future studies.

## Conclusions

In conclusion, major differences in economic burden between BrS and LQTS patients were identified in this study. These findings can offer novel insight into the financial management of clinical interventions and optimization of healthcare policies surrounding BrS and LQTS. However, it is imperative that further research is conducted to extend the costs analysis amongst subgroups in both cohorts.

## Data Availability

All data produced in the present study are available upon reasonable request to the authors

## Conflict of interest

None.

## Funding

None.

## Acknowledgements

None.

## Author Contributions

SL and CTC—statistical analysis, data analysis, data interpretation, cost analysis, manuscript drafting;

DR, OHIC, TTLL, KSKL, LR, RR, GB, KPL, SZ, FZL, WTW, TL —data analysis, manuscript revision

GT—data acquisition, database building, cost analysis, study conception, statistical analysis, manuscript drafting, manuscript revision

## Notes

### Competing Interest Statement

The authors have declared no competing interest.

### Funding Statement

This study was funded by a Clinical Assistant Professorship at the Chinese University of Hong Kong

### Author Declarations

The study was part of a wider study on cardiac arrhythmias approved by The Joint Chinese University of Hong Kong-New Territories East Cluster Clinical Research Ethics Committee.

## References

1. Lazzerini PE, Capecchi PL, El-Sherif N, Laghi-Pasini F, Boutjdir M. Emerging Arrhythmic Risk of Autoimmune and Inflammatory Cardiac Channelopathies. J Am Heart Assoc. 2018;7(7):e010595.

2. Ortiz-Bonnin B, Rinné S, Moss R, Streit AK, Scharf M, Richter K, et al. Electrophysiological characterization of a large set of novel variants in the SCN5A-gene: identification of novel LQTS3 and BrS mutations. Pflugers Arch. 2016;468(468):1375–87.

3. Aiba T. Recent understanding of clinical sequencing and gene-based risk stratification in inherited primary arrhythmia syndrome. J Cardiol. 2019;73(73):335–42.

4. Snir AD, Raju H. Current Controversies and Challenges in Brugada Syndrome. Eur Cardiol. 2019;14(14):169–74.

5. Letsas KP, Asvestas D, Baranchuk A, Liu T, Georgopoulos S, Efremidis M, et al. Prognosis, risk stratification, and management of asymptomatic individuals with Brugada syndrome: A systematic review. Pacing Clin Electrophysiol. 2017;40(40):1332–45.

6. Aziz HM, Zarzecki MP, Garcia-Zamora S, Kim MS, Bijak P, Tse G, et al. Pathogenesis and Management of Brugada Syndrome: Recent Advances and Protocol for Umbrella Reviews of Meta-Analyses in Major Arrhythmic Events Risk Stratification. J Clin Med. 2022;11(7).

7. Chung CT, Bazoukis G, Radford D, Coakley-Youngs E, Rajan R, Matusik PT, et al. Predictive risk models for forecasting arrhythmic outcomes in Brugada syndrome: A focused review. J Electrocardiol. 2022;72:28–34.

8. Boriani G, Cimaglia P, Biffi M, Martignani C, Ziacchi M, Valzania C, et al. Cost-effectiveness of implantable cardioverter-defibrillator in today’s world. Indian Heart J. 2014;66 Suppl 1(Suppl 1):S101–4.

9. Zhang M, Ren Y, Wang L, Jia J, Tian L. Cost-Effectiveness of Dronedarone and Amiodarone for the Treatment of Chinese Patients With Atrial Fibrillation. Front Public Health. 2021;9:726294.

10. Chung CT, Lee S, Zhou J, Chou OHI, Lee TTL, Leung KSK, et al. Clinical Characteristics, Genetic Basis and Healthcare Resource Utilisation and Costs in Patients with Catecholaminergic Polymorphic Ventricular Tachycardia: A Retrospective Cohort Study. Reviews in Cardiovascular Medicine. 2022;23(23):276.

11. StreptomycesChan JSK, Lakhani I, Lee TTL, Chou OHI, Lee YHA, Cheung YM, et al. Cardiovascular Outcomes and Hospitalizations in Asian Patients Receiving Immune Checkpoint Inhibitors: A Population-based Study. Curr Probl Cardiol. 2022;48(48):101380.

12. Wong JYH, Luk LYF, Yip TF, Lee TTL, Wai AKC, Ho JWK. Incidence of Emergency Department Visits for Sexual Abuse Among Youth in Hong Kong Before and During the COVID-19 Pandemic. JAMA Netw Open. 2022;5(5):e2236278.

13. Wai AK, Chan CY, Cheung AW, Wang K, Chan SC, Lee TT, et al. Association of Molnupiravir and Nirmatrelvir-Ritonavir with preventable mortality, hospital admissions and related avoidable healthcare system cost among high-risk patients with mild to moderate COVID-19. Lancet Reg Health West Pac. 2022:100602.

14. Al-Khatib SM, Stevenson WG, Ackerman MJ, Bryant WJ, Callans DJ, Curtis AB, et al. 2017 AHA/ACC/HRS Guideline for Management of Patients With Ventricular Arrhythmias and the Prevention of Sudden Cardiac Death: A Report of the American College of Cardiology/American Heart Association Task Force on Clinical Practice Guidelines and the Heart Rhythm Society. J Am Coll Cardiol. 2018;72(72):e91–e220.

15. Lee S, Zhou J, Chung CT, Lee ROY, Bazoukis G, Letsas KP, et al. Comparing the Performance of Published Risk Scores in Brugada Syndrome: A Multi-center Cohort Study. Curr Probl Cardiol. 2022;47(47):101381.

16. Lee S, Zhou J, Jeevaratnam K, Wong WT, Wong ICK, Mak C, et al. Paediatric/young versus adult patients with long QT syndrome. Open Heart. 2021;8(2).

17. Tse G, Lee S, Zhou J, Liu T, Wong ICK, Mak C, et al. Territory-Wide Chinese Cohort of Long QT Syndrome: Random Survival Forest and Cox Analyses. Front Cardiovasc Med. 2021;8:608592.

18. Lee S, Zhou J, Li KHC, Leung KSK, Lakhani I, Liu T, et al. Territory-wide cohort study of Brugada syndrome in Hong Kong: predictors of long-term outcomes using random survival forests and non-negative matrix factorisation. Open Heart. 2021;8(1).

19. Ruiter JS, Berkenbosch-Nieuwhof K, van den Berg MP, van Dijk R, Middel B, van Tintelen JP. The importance of the family history in caring for families with long QT syndrome and dilated cardiomyopathy. Am J Med Genet A. 2010;152a(3):607–12.

20. Phillips KA, Ackerman MJ, Sakowski J, Berul CI. Cost-effectiveness analysis of genetic testing for familial long QT syndrome in symptomatic index cases. Heart Rhythm. 2005;2(2):1294–300.

21. Waddell-Smith KE, Skinner JR. Update on the Diagnosis and Management of Familial Long QT Syndrome. Heart Lung Circ. 2016;25(25):769–76.

22. Angrist M, Chandrasekharan S, Heaney C, Cook-Deegan R. Impact of gene patents and licensing practices on access to genetic testing for long QT syndrome. Genet Med. 2010;12(4 Suppl):S111–54.

23. Gnecchi M, Sala L, Schwartz PJ. Precision Medicine and cardiac channelopathies: when dreams meet reality. Eur Heart J. 2021;42(42):1661–75.

24. Perez MV, Kumarasamy NA, Owens DK, Wang PJ, Hlatky MA. Cost-effectiveness of genetic testing in family members of patients with long-QT syndrome. Circ Cardiovasc Qual Outcomes. 2011;4(4):76–84.

25. Itoh H, Crotti L, Aiba T, Spazzolini C, Denjoy I, Fressart V, et al. The genetics underlying acquired long QT syndrome: impact for genetic screening. Eur Heart J. 2016;37(37):1456–64.

26. Hermans BJM, Bennis FC, Vink AS, Koopsen T, Lyon A, Wilde AAM, et al. Improving long QT syndrome diagnosis by a polynomial-based T-wave morphology characterization. Heart Rhythm. 2020;17(5 Pt A):752–8.

